# High proportion of tuberculosis transmission among social contacts in rural China: a 12-year prospective population-based genomic epidemiological study

**DOI:** 10.1101/2022.04.18.22273772

**Authors:** Meng Li, Mingcheng Guo, Ying Peng, Qi Jiang, Lan Xia, Sheng Zhong, Yong Qiu, Xin Su, Shu Zhang, Chongguang Yang, Peierdun Mijiti, Qizhi Mao, Howard Takiff, Fabin Li, Chuang Chen, Qian Gao

**Author notes:** **Correspondence to:** Dr. Qian Gao, Key Laboratory of Medical Molecular Virology (MOE/NHC/CAMS), School of Basic Medical Science, Shanghai Medical College, Shanghai Institute of Infectious Disease and Biosecurity, Fudan University, Shanghai, China, 131 Dongan Road, Shanghai, China 200032, Chuang Chen, Institution for Tuberculosis Prevention and Control, Sichuan Provincial Center for Disease Control and Prevention, 6 Zhongxue Road, Chengdu, China 610041, Fabin Li, Heilongjiang Provincial Center for Tuberculosis Prevention and Control, 40 Youfang Road, Harbin, China 150036. These authors contributed equally to this study (Meng Li, Mingcheng Guo, Ying Peng). These authors contributed equally to this study (Fabin Li, Chuang Chen, Qian Gao).

## Abstract

**Background:** Tuberculosis (TB) is more prevalent in rural than urban areas in China, and delineating TB transmission patterns in rural populations could improve TB control.

**Methods:** We conducted a prospective population-based study of culture-positive pulmonary TB patients diagnosed between July 1, 2009 and December 31, 2020 in two rural counties in China. Genomic clusters were defined with a threshold distance of 12-single-nucleotide-polymorphisms, based on whole-genome sequencing. Risk factors for clustering were identified by logistic regression. Transmission links were sought through epidemiological investigation of genomic-clustered patients.

**Findings:** Of 1517 and 751 culture-positive pulmonary TB patients in Wusheng and Wuchang counties, respectively, 1289 and 699 strains were sequenced. Overall, 624 (31·4%, 624/1988) patients were grouped into 225 genomic clusters. Epidemiological links were confirmed in 41·8% (196/469) of clustered isolates, including family (32·7%, 64/196) and social contacts (67·3%, 132/196). Social contacts were generally with relatives, within the community or in shared aggregated settings outside the community, but the proportion of clustered contacts in each category differed between the two sites. The time interval between diagnosis of student cases and contacts was significantly shorter than family and social contacts, probably due to enhanced student contact screening. Transmission of multidrug-resistant strains was likely responsible for 81·4% (83/102) of MDR-TB cases, with minimal acquisition of additional resistance mutations.

**Interpretation:** A large proportion of TB transmission in rural China occurred among social contacts, suggesting that active screening and aggressive contact tracing could benefit TB control, but contact screening should be tailored to local patterns of social interactions.

**Funding:** National Science and Technology Major Project of China, Natural Science Foundation of China, and Science and Technology Major Project of Shanghai

**Evidence before this study:** We searched PubMed for genomic epidemiological studies of *Mycobacterium tuberculosis* published in English before April 2022 employing whole-genome sequencing, using the search terms “tuberculosis”, “transmission”, “population based”, and “whole-genome sequencing”. We identified only 11 studies in which whole-genome sequencing was used to investigate transmission of *M tuberculosis* at the population level. We also searched the China national knowledge infrastructure (CNKI) and WANFANG databases with the same search terms for papers published in Chinese, but did not identify any studies. The duration of most of the 11 studies we identified was less than 5 years. Seven studies conducted epidemiological investigations of genomic-clustered cases, but the proportion of cases with confirmed epidemiological links was very low. Therefore, no studies had sufficient evidence to identify populations and sites at high risk of TB transmission. Five studies were conducted in China but all were in urban areas and focused on MDR-TB patients and internal migrants. The pattern of TB transmission in rural China, where TB is more prevalent, had not been addressed.

**Added value of this study:** To our knowledge, ours is the first population-based genomic epidemiological study to delineate TB transmission patterns in rural China. Close contacts have been shown to be a high-risk group for TB transmission in other countries. In China, however, the huge number of TB patients, limited resources for TB prevention and control and the stigma associated with tuberculosis all contribute to a failure to identify and screen many close contacts. As a consequence, close contacts have been calculated to contribute only about 2% of the total TB burden. In this study, through the investigation of genomic-clustered patients, we found at least 41·8% of clustered patients were close contacts who comprised 9·9% of the total TB patients in the study. Moreover, more than two-thirds of the close contacts were social contacts rather than members of the immediate family. The composition of social contacts differed between the two study sites due to differences in climate and lifestyle habits. The average time interval between the diagnosis of clustered student contacts was shorter than for family or community contacts. In addition, transmission of MDR strains was likely responsible for 81·4% of MDR-TB cases, with minimal acquisition of additional resistance mutations. Our 12-year study identified patterns of TB transmission not identified by previous studies, demonstrating the value of long-term genomic epidemiological studies.

**Implications of all the available evidence:** Our study demonstrates that much of the transmission of TB in rural China was among close contacts, especially social contacts. Therefore, strengthening and improving proactive screening of close social contacts can identify more TB patients and shorten the time to patient detection. We believe that this type of vigorous active case-finding is essential for reducing TB transmission and the considerable TB burden in China. Long-term prospective genomic epidemiological studies provide a useful picture of TB transmission patterns that can help guide the design of strategies to improve TB prevention and control.

## Introduction

Tuberculosis (TB) remains an important threat to global health, with an estimated 10 million people worldwide falling ill and at least 1·4 million deaths in 2019.^1^ Although the global incidence of TB has declined in recent years, there is still a significant gap between current rates of decline and the goals of the WHO End TB Strategy.^2,3^

It is estimated that approximately one-third of TB patients worldwide were not diagnosed,^1^ so measures that increase diagnosis, especially early in the disease, are key to reducing transmission and thereby controlling TB. One of the core strategies to increase TB diagnosis and achieve global TB control is active case-finding.^4^ The World Health Organization (WHO) recommends systematic TB screening of the general population in areas with an estimated TB prevalence of 0·5% or higher.^5^ For areas with a lower TB prevalence, a cost-effective strategy involves identifying populations at high risk for transmission and then implementing targeted screening.

Genomic-epidemiological studies of tuberculosis can help identify populations at high risk of transmission, but the limited discriminatory power of previously employed genotyping methods and the barriers to epidemiological investigations have made it difficult to define these populations in China.^6^ The populations at high risk for TB in other countries, such as HIV-positive, homeless, and drug abusing individuals, are less important contributors to the TB burden in China.^5,7^ Instead, TB screening in China is focused principally on the elderly and internal migrants, but because of the large size of these populations in China, generalized screening is not practical. Most genomic-epidemiological studies of tuberculosis in China have been conducted in large cities, but China also has extensive rural areas where the prevalence of TB is up to three times higher than in urban regions.^8^ Therefore, identifying high-risk populations and delineating transmission patterns in rural areas is important for improving TB control in China. To define the transmission patterns of the rural TB burden and thereby provide guidance for improving TB control, we conducted a 12-year prospective population-based genomic-epidemiological study in two rural counties in China.

## Methods

### Study design and participants

Wusheng and Wuchang are rural counties in southwestern and northeastern China, respectively. Wusheng is located in the east of Sichuan Province, with an area of 960 square kilometers and an estimated 556,000 inhabitants in 2020. Wuchang is located in the south of Heilongjiang Province, with an area of 7,512 square kilometers. The study included 14 townships (2,299·5 square kilometers) in Wuchang, with a combined estimated 419,000 inhabitants in 2020. The average annual reported TB incidences were 95·8 (per 100,000 population) in Wusheng and 68·9 in Wuchang. Lower incidences recorded in 2020 were probably inaccurate and a result of reduced TB diagnosis due to the COVID-19 pandemic.

The study population was comprised of all culture-positive pulmonary TB patients 15 years or older who were diagnosed between July 1, 2009 and December 31, 2020 in the regions included in the study. The study was approved by the institutional review board of the Institutes of Biomedical Sciences, Fudan University and all enrolled patients provided written informed consent.

### Diagnostic procedures

The Wusheng County Center for Disease Control and Prevention and the Wuchang City Center for Tuberculosis Control and Prevention are responsible for the diagnosis and treatment of TB in their respective areas. Individuals with TB-like symptoms or abnormal chest radiographs in general hospitals and township health centers are referred to these designated medical institutions for diagnosis by sputum smear and culture. Sputum specimens were collected when the patients presented for TB diagnosis, before starting treatment, and culture-positive isolates were stored in the site laboratory.

### Whole-genome sequencing

All stored clinical strains isolated during the study period were re-cultured from frozen stocks. DNA was extracted using the CTAB method and sequenced as previously described.^9^ Raw sequence reads were trimmed with Sickle (version 1·33) and aligned to the inferred *Mycobacterium tuberculosis* complex ancestor sequence^10^ using BWA-MEM. SAMtools (version 1·3·1) and Varscan (version 2·3·6) were then used to identify single nucleotide polymorphisms (SNPs). Pairwise SNP distances were calculated based on fixed SNPs (frequency ≥75%), excluding those in drug-resistance associated genes and repetitive regions of the genome (e.g., PPE/PE-PGRS family genes, phage sequences, and insertion or mobile genetic elements). A genomic cluster was defined as strains differing by 12 or fewer SNPs, consistent with linkage through recent transmission.^11^ Sequencing data were deposited in the Genome Sequence Archive (https://bigd.big.ac.cn/gsa) under BioProject PRJCA008815 and PRJCA008816.

Based on the identified SNPs, a phylogeny tree was constructed with RAxML-NG (version 1·0·2) software, using the maximum likelihood method with 100 bootstraps and visualized with Interactive Tree of Life (https://itol.embl.de/). Strains were classified into different lineages according to Coll et al.^12^ Strains belonging to lineage 2 (L2), also termed the Beijing family, were divided into L2·3, representing “modern” Beijing, and other sub-lineages considered “ancient” Beijing.^13^

Drug-resistance profiles were predicted for 14 anti-TB drugs based on the mutations reported to be associated with resistance,^14^ using Whole-genome sequencing (WGS) data as described previously.^15^ Pan-susceptible TB was defined as susceptibility to the four first-line drugs (isoniazid, rifampicin, ethambutol and pyrazinamide). Multidrug-resistant TB (MDR-TB) was defined as resistance to at least isoniazid and rifampicin. Strains with resistance to any of the four first line drugs but not MDR were termed other drug resistant (DR) TB.

### Epidemiological investigation

We conducted epidemiological investigations twice. First, at the time of TB diagnosis we collected demographic, clinical and laboratory information on each patient and also collected data on their close contacts. We then conducted in-depth epidemiological investigation of patients whose strains belonged to genomic clusters, including their residences, workplaces, and public community centers and facilities frequented in the three years prior to their TB diagnosis. Putative transmission networks were constructed based on the structure of the genomic phylogeny and the epidemiologic links. The epidemiologic links were defined as confirmed -- patients who knew each other and had a history of contact before diagnosis, or probable -- patients who did not know each other but lived in the same village.

### Statistical analysis

The clustering rate was calculated by the N method and the cumulative clustering rate was obtained by calculating the clustering rate in 2009 and then adding the TB patients from every successive year of the study, as previously described.^16^ Changes in temporal trends of annual clustering were detected using joinpoint regression analysis. The distribution of continuous and categorical variables between groups was compared using the Wilcoxon rank sum test or the chi-square test. Logistic regression analysis was used to calculate the odds ratios (OR) and 95% confidence intervals (CI) for risk factors associated with genomic clustering. Variables with p-values less than 0·2 in the univariable analysis were included in the multivariable analysis to calculate the adjusted odds ratios (aOR). Factors with a p-value less than 0·05 in the final model were considered statistically significant. All analyses were performed in Stata (version 14·0).

## Results

Between July 1, 2009 and December 31, 2020, 5502 and 3435 pulmonary TB patients were officially registered in Wusheng and Wuchang, respectively, of whom 1517 and 751 had positive sputum cultures. After excluding 103 strains that failed in re-culture, 1452 and 713 strains were sequenced. We further excluded 123 patients with non-tuberculous mycobacteria isolates and 54 patients with recurrent TB. The final analysis thus included 1289 and 699 patients from Wusheng and Wuchang, respectively (Figure 1), whose characteristics are shown in Table 1. The proportions of patients less than 25 years or students were higher in Wusheng [15·4% (198/1289) and 7·6% (98/1289)] than in Wuchang [9·9% (69/699) and 2·9% (20/699)]. Patients in Wuchang were more likely to have had longer diagnosis delays, chest cavities and smear positivity.

**Figure 1:**
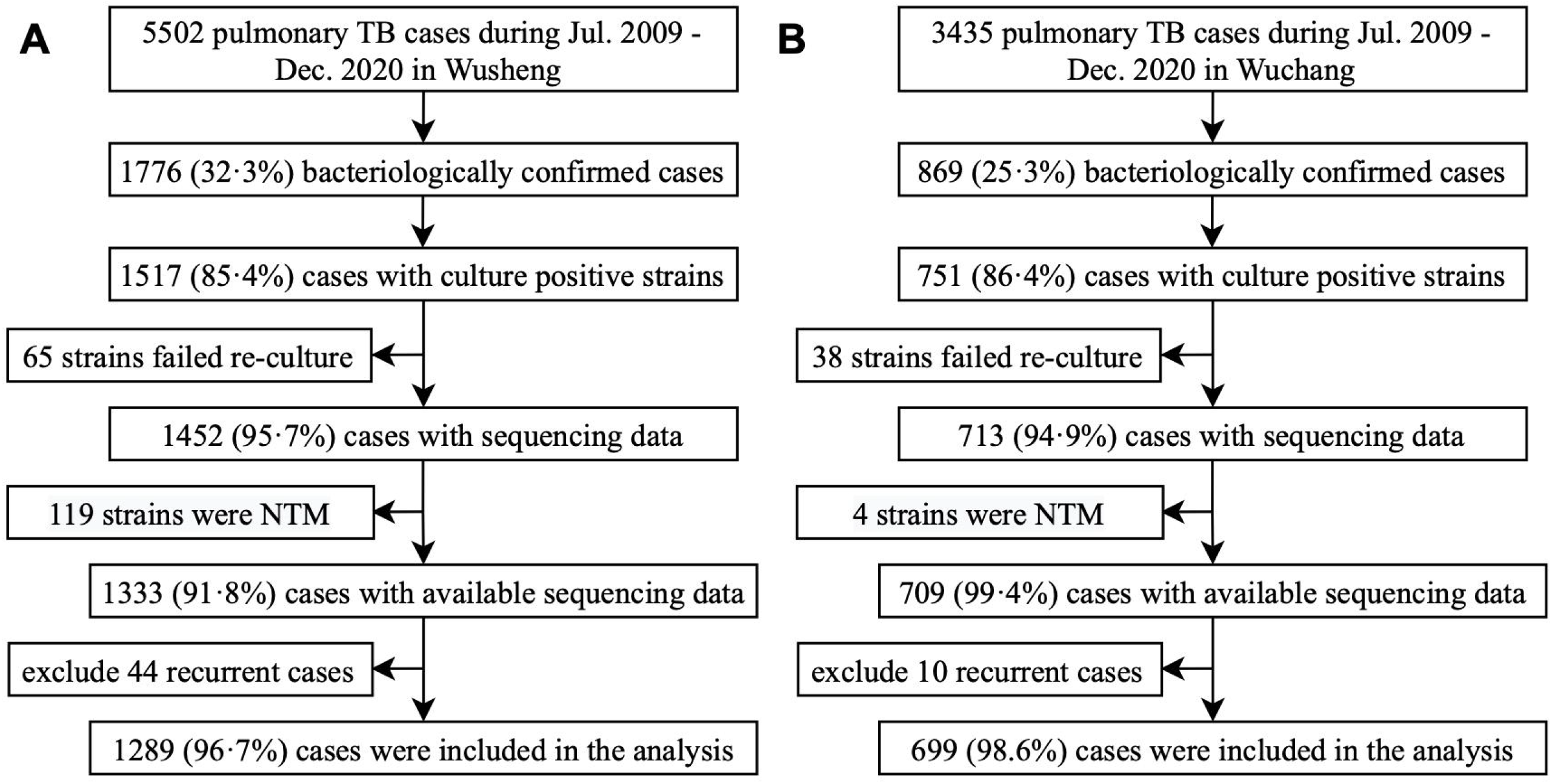
Sample enrollment in Wusheng (A) and Wuchang (B). NTM=non-tuberculous mycobacteria.

**Table 1:** Characteristics of tuberculosis patients in Wusheng and Wuchang, 2009-2020.

|  | Wusheng (n=1289) | Wuchang (n=699) |
| --- | --- | --- |
| Sex |  |  |
| Female | 271 (21.0) | 202 (28.9) |
| Male | 1018 (79.0) | 497 (71.1) |
| Age |  |  |
| <25 | 198 (15.4) | 69 (9.9) |
| 25–44 | 328 (25.4) | 193 (27.6) |
| 45–64 | 519 (40.3) | 296 (42.3) |
| ≥65 | 244 (18.9) | 141 (20.2) |
| Occupation |  |  |
| Farmer | 1096 (85.0) | 578 (82.7) |
| Students | 98 (7.6) | 20 (2.9) |
| Others | 95 (7.4) | 101 (14.4) |
| History of tuberculosis |  |  |
| New | 1197 (92.9) | 642 (91.8) |
| Retreated | 92 (7.1) | 57 (8.2) |
| Diagnostic delay |  |  |
| <2 weeks | 428 (33.2) | 103 (14.7) |
| 2–4 weeks | 227 (17.6) | 162 (23.2) |
| 4–8 weeks | 371 (28.8) | 208 (29.8) |
| ≥8 weeks | 263 (20.4) | 226 (32.3) |
| Chest cavitation |  |  |
| No | 889 (69.0) | 407 (58.2) |
| Yes | 400 (31.0) | 292 (41.8) |
| Sputum smear status |  |  |
| Negative | 634 (49.2) | 244 (34.9) |
| Positive | 655 (50.8) | 455 (65.1) |
| Drug-resistance profile |  |  |
| Pan-susceptible | 1128 (87.5) | 588 (84.1) |
| Other DR | 95 (7.4) | 75 (10.7) |
| MDR | 66 (5.1) | 36 (5.2) |

Data are n (%) unless otherwise specified. DR=drug resistance. MDR=multidrug resistance.

Phylogenetic analysis revealed that most strains belonged to the Beijing family lineage (69·0%, 1372/1988) (Figure 2), with more belonging to the modern Beijing sublineage (74·1%, 1016/1372) than ancient Beijing sublineages (25·9%, 356/1372). All of the non-Beijing strains belonged to L4 (99·8%, 615/616), except for one L1 strain. Analysis of the genome sequences for mutations conferring resistance to 14 anti-TB drugs showed that the drug-resistance profiles were similar between the two sites (Figure 2; Table 2). In total, there were 1716 (86·3%) pan-susceptible strains and 272 (13·7%) strains resistant to at least one anti-TB drug. The isoniazid-resistant, rifampicin-resistant and MDR strains accounted for 10·7% (212/1988), 7·6% (152/1988) and 5·1% (102/1988) respectively, and 29·4% (30/102) of MDR strains were predicted to also be resistant to fluoroquinolones.

**Figure 2:**
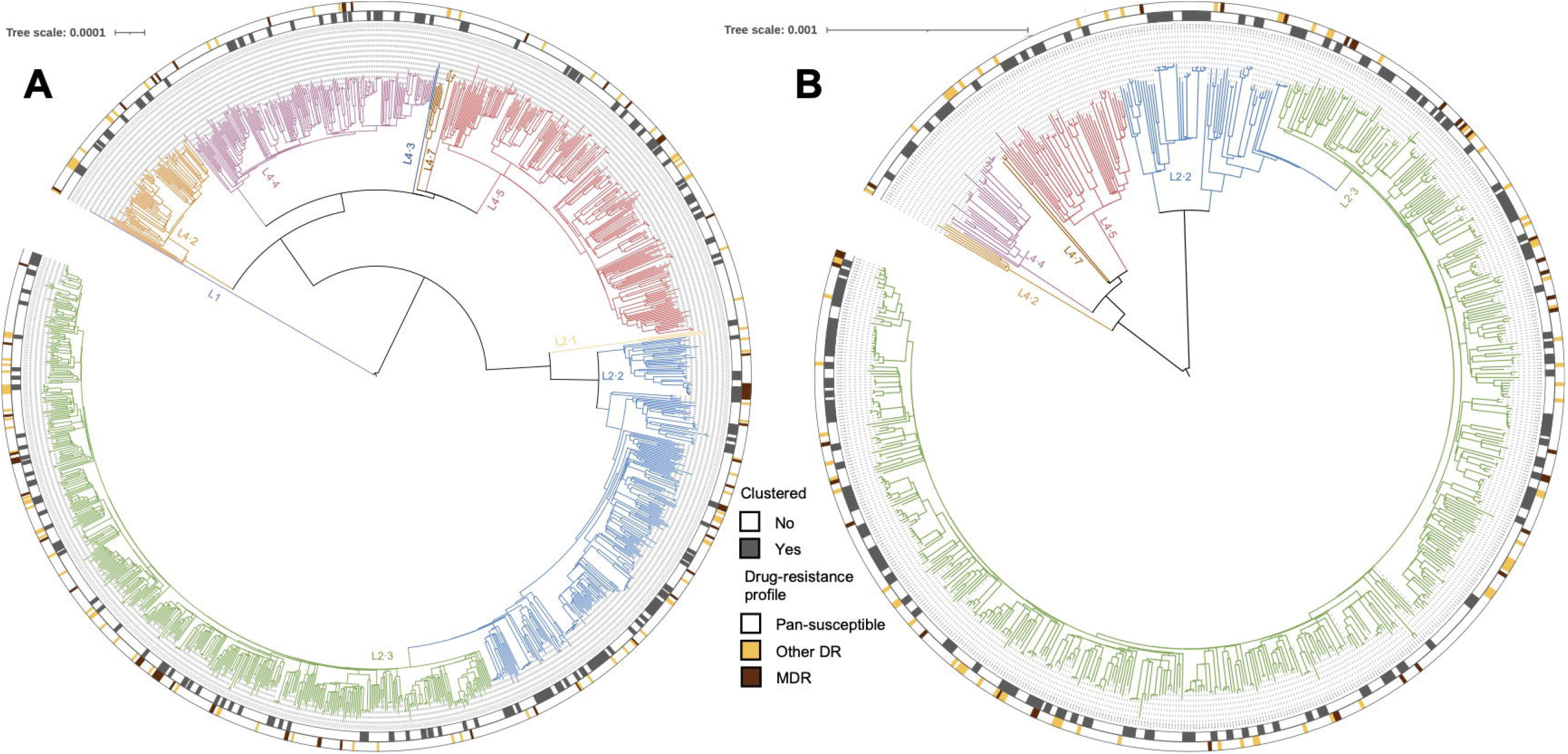
Phylogeny, clustering, and resistance profile of 1289 *Mycobacterium tuberculosis* strains isolated in Wusheng (A) and 699 *Mycobacterium tuberculosis* strains isolated in Wuchang (B). The different colors on the branches indicate different lineages and sublineages. The outer grey circle indicates genomic-clustered strains differing by ≤ 12 single-nucleotide polymorphisms. The outer yellow-red circle indicates other drug resistance and multidrug resistance.

**Table 2:** Drug-resistance profile, stratified by new and retreated cases.

|  | New cases (n=1839) | Retreated cases (n=149) | Total (n=1988) |
| --- | --- | --- | --- |
| Pan-susceptible | 1610 (87.5) | 106 (71.1) | 1716 (86.3) |
| Resistance to INH | 180 (9.8) | 32 (21.5) | 212 (10.7) |
| Resistance to RIF | 119 (6.5) | 33 (22.1) | 152 (7.6) |
| Resistance to EMB | 54 (2.9) | 22 (14.8) | 76 (3.8) |
| Resistance to PZA | 24 (1.3) | 9 (6.0) | 33 (1.7) |
| MDR | 78 (4.2) | 24 (16.1) | 102 (5.1) |
| MDR plus resistance to FQ | 24 (1.3) | 6 (4.0) | 30 (1.5) |

Data are n (%) unless otherwise specified. INH=isoniazid. RIF=rifampicin. EMB=ethambutol.
PZA=pyrazinamide. MDR=multidrug resistance. FQ=fluoroquinolone.

To estimate the level of recent transmission, we calculated the clustering rate between 2009 to 2020. A total of 624 (31·4%, 624/1988) strains, 347 (26·9%, 347/1289) from Wusheng and 277 (39·6%, 277/699) from Wuchang, were grouped into 225 genomic clusters containing 2 to 13 strains (Figure 2). To understand the dynamics of clustering trends in the two sites, we calculated the cumulative clustering rate and found that the rate gradually increased as more strains were analyzed, but the trend eased between 2015 and 2016 in both sites. As seen in Figure 3, the clustering rate increased significantly after 2016, especially in Wusheng (p=0·004, Figure S1, rising from 21·8% in 2017 to 26·9% in 2020), which may have been the result of strategies we implemented starting in 2017: active case-finding based on symptoms identified 15-25% more patients; stepwise sputum collection to improve the diagnostic quality of the sputum^17^ and sputum cultures in liquid media increased the percentage of culture-positive patients from 20-30% to 40-50%.

**Figure 3:**
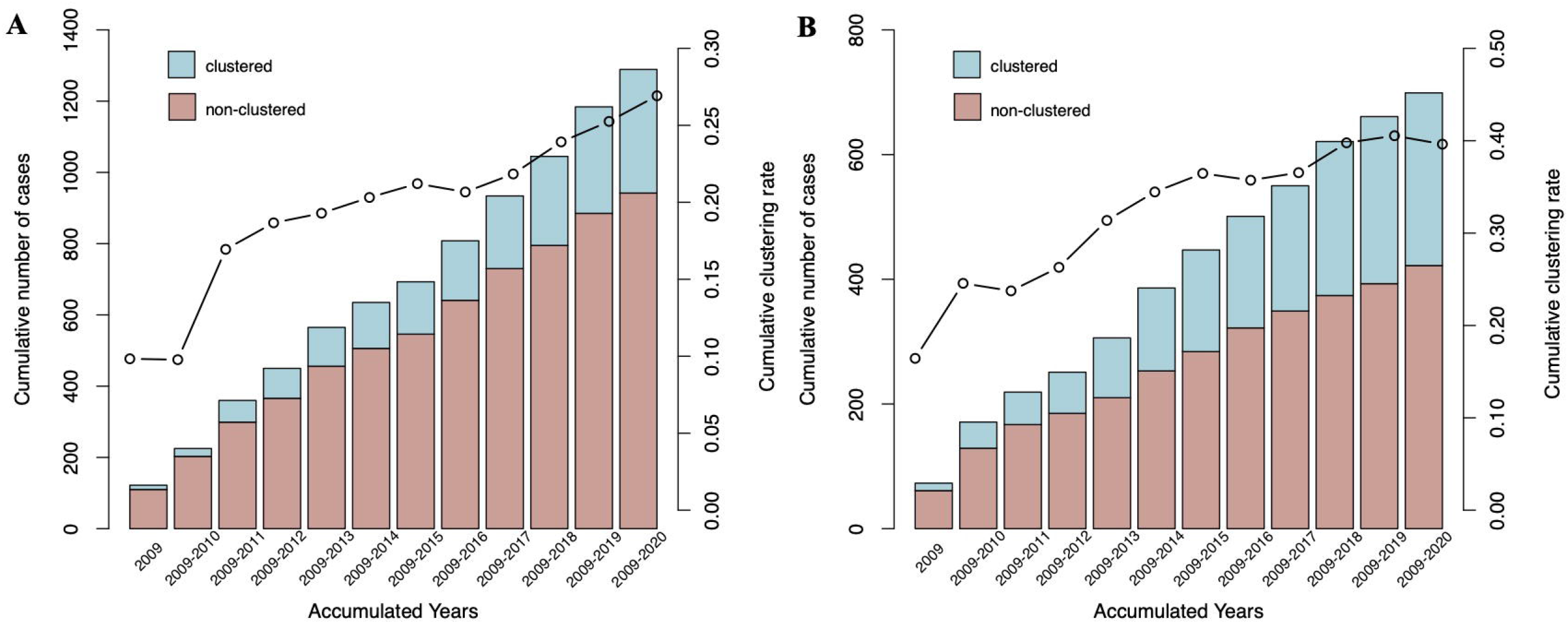
Cumulative clustering rate in Wusheng (A) and Wuchang (B), 2009-2020. The bar indicates the cumulative number of cases, the line indicates the cumulative clustering rate.

In the analysis of risk factors for MDR-TB, the only association was a higher proportion of MDR in retreated patients (16·1%, 24/149) than in new cases (4·2%, 78/1839). To investigate the cause of MDR we analyzed the number of clustered and non-clustered patients in the 102 MDR patients and found that 34 were clustered. While retreated MDR-TB cases could have secondary resistance that was acquired during treatment, new cases were presumably infected with MDR strains and represent primary resistance. Among the non-clustered MDR patients, 49 were new patients. Therefore, in total, 81·4% (83/102) of MDR-TB patients were likely caused by the transmission of MDR strains. We also looked for accumulated additional drug-resistance mutations along the transmission chain. Among 32 transmission events of MDR-TB belonging to 11 genomic-clusters, we found just two events (6·3%, 2/32) of accumulation of additional resistance (Figure S2), which is a lower incidence of acquired resistance than has been reported in urban areas of China.^11^

To further delineate the transmission links between the genomic-clustered patients we performed in-depth epidemiological investigation of all patients with clustered isolates. These investigations were completed for 469 (75·2%, 469/624) patients, 294 in Wusheng and 175 in Wuchang, but the other 155 clustered patients (24·8%) had either died or were lost to follow-up. Confirmed epidemiological links were identified in 196 (41·8%, 196/469) of the genomic-clustered patients investigated (110 in Wusheng and 86 in Wuchang), and probable epidemiological links were identified in 26 (5·5%, 26/469) patients. The characteristics of each cluster with confirmed or probable epidemiological links are shown in Table S1.

The 196 patients with confirmed epidemiological links were all close contacts. Among them (Figure 4A), transmission between family contacts was identified in 32·7% (64/196) and between social contacts in 67·3% (132/196). To further analyze transmission patterns, we divided the social contacts into three categories: relatives outside of the immediate family, community contacts, and contacts who shared aggregated settings such as teahouses located outside the community. Epidemiological links were found in community contacts in 14·5% (16/110) of the confirmed transmission in Wusheng but 51·2% (44/86) in Wuchang. Surprisingly, the proportions were reversed for confirmed epidemiological links in aggregated settings outside the community, with 43·6% (48/110) in Wusheng and 15·1% (13/86) in Wuchang. The percentages of confirmed links to relatives beyond the immediate family were low in both sites, with 4·5% (5/110) in Wusheng and 7·0% (6/86) in Wuchang.

**Figure 4:**
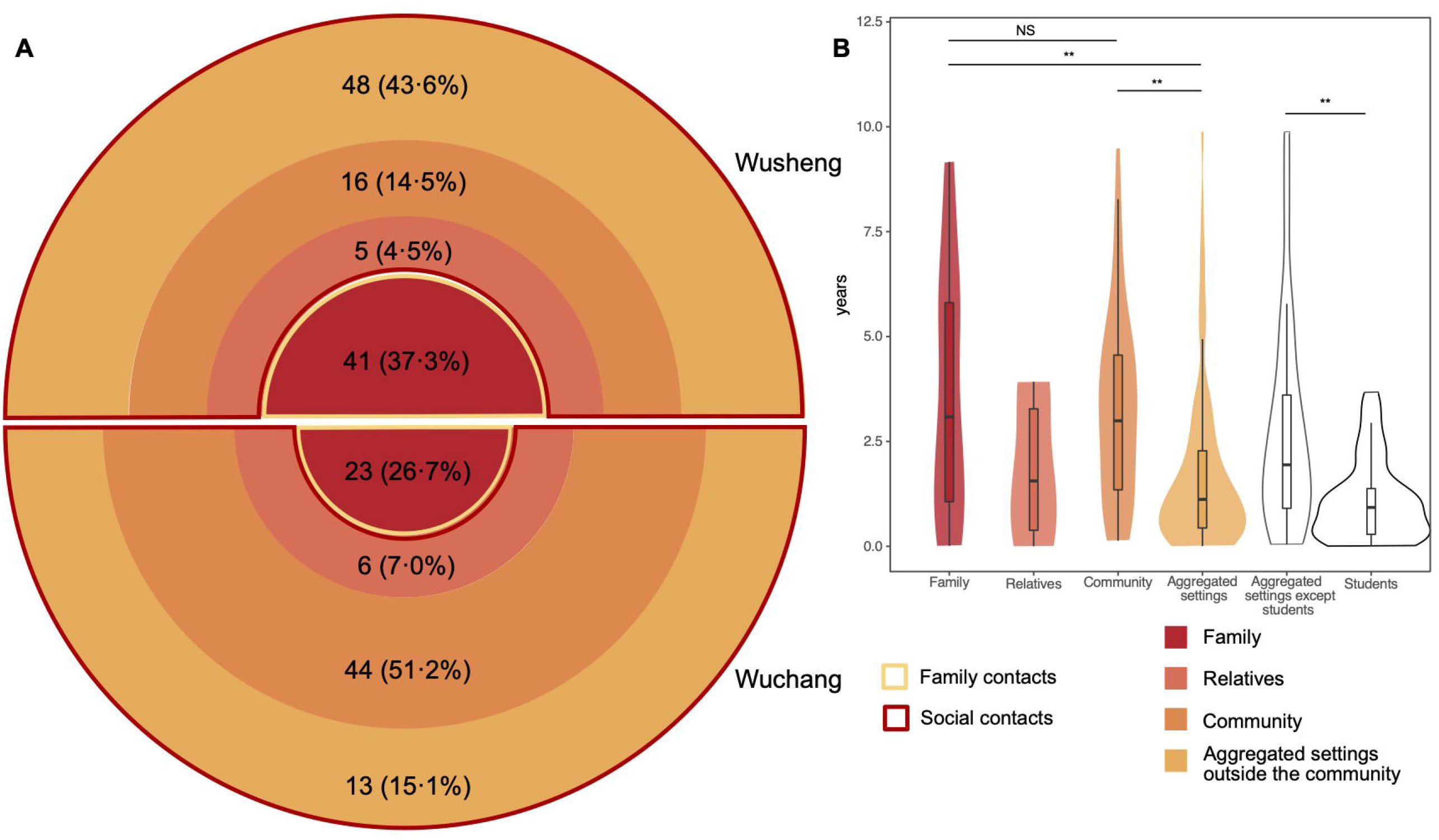
Categories of confirmed epidemiological links and their time interval between diagnoses. The four colors of the circle indicate four categories of contacts with confirmed epidemiological links. The darker the color, the closer the relationship. (A) The yellow border indicates family contacts (Family) and the red border indicates social contacts (Relatives, Community and Aggregated settings outside the community). The number in the ring indicates the number and proportion of each category of contacts. The width of the ring indicates the proportion of each category. (B) The time interval between diagnoses in the four categories of contacts and students. Due to the small number of relatives, they were not compared with the other categories. ^**^p<0.01, NS refers to no significance (given by Wilcoxon non-parametric rank sum test).

We constructed putative transmission networks based on genomic phylogeny and epidemiological links. Figure 5 shows examples of three of the largest clusters. The inhabitants of the villages in Wuchang live in fairly close proximity, often with daily contact in village shops that may explain the high level of transmission occurring within the community (Figure 5A). In Wusheng, by contrast, the majority of identified epidemiological links were outside the community and mainly in schools and teahouses (Figure 5B/5C). The teahouses in Wusheng are generally located in townships outside the villages and are frequented by people from several different villages.

**Figure 5:**
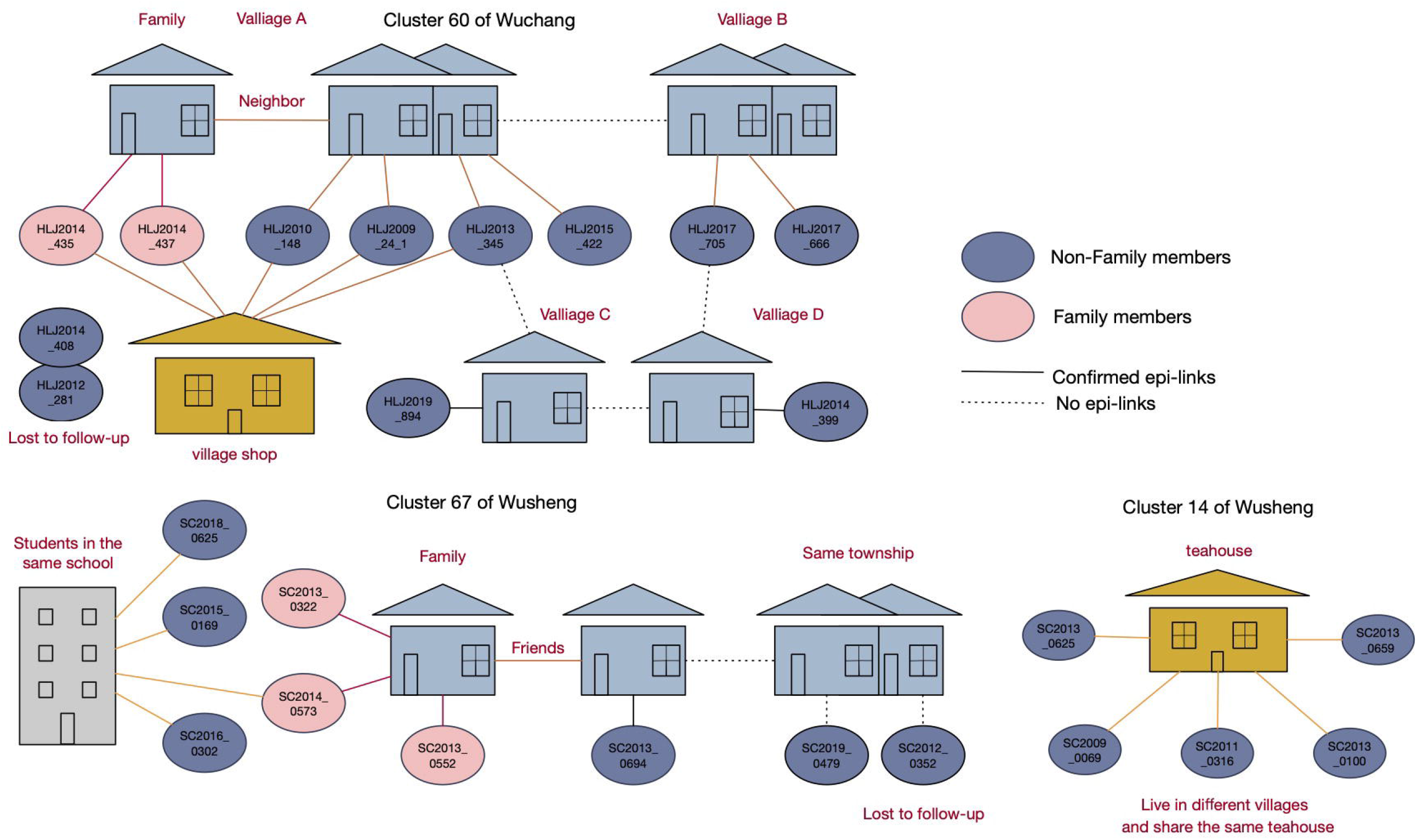
Putative transmission network for 3 clusters based on the structure of the genomic phylogeny and the epidemiological links. The color of the line indicates the categories of contacts in Figure 4. epi-link=epidemiological link.

We then analyzed the time interval between TB diagnosis in clustered cases in the different categories of contacts (Figure 4B). We found no statistical difference (p=0·90) in the time interval between diagnosis of family contacts (3·1 years, Interquartile range [IQR] 0·9-5·9 years) and community contacts (3·0 years, IQR 1·3-4·6 years). Unexpectedly though, the average diagnostic time interval between contacts who shared the aggregated settings outside the community (1·1 years, IQR 0·4-2·3 years) was significantly shorter than for family (p<0·01) or community contacts (p<0·01). This shorter interval was perhaps because most of the contacts who shared aggregated settings outside the community were students, and the average time interval between the diagnosis of student contacts (0·9 years, IQR 0·3-1·4 years) was significantly shorter (p<0·01) than for all other contacts (1·9 years, IQR 0·7-3·6 years).

Finally, we used logistic regression to identify risk factors associated with clustering. The univariate analysis found that occupation and the Beijing strain were significantly associated with clustering, and both associations persisted in the multivariate analysis (Table 3). Patients with a Beijing lineage strain had a greater risk of clustering (aOR 1·46, 95% CI, 1·17-1·82, p*=*0·001) than patients with L4 strains. Also, compared to any other occupation, farmers had a lower risk (aOR 0·60, 95% CI, 0·44-0·82, p=0·001) and students had a higher risk (aOR, 2·11; 95% CI, 1·20-3·71, p=0·01) of clustering.

**Table 3:** Univariate and multivariable logistic regression of risk factors for clustering.

|  | Clustered<br>(n=624) | Non-clustered<br>(n=1364) | Univariate regression |  | Multivariable regression |  |
| --- | --- | --- | --- | --- | --- | --- |
|  |  |  | OR (95% CI) | p value | aOR (95% CI) | p value |
| Sex |  |  |  |  |  |  |
| Female | 148 (31.3) | 325 (68.7) | 1.00 |  | .. |  |
| Male | 476 (31.4) | 746 (68.6) | 1.01 (0.81, 1.26) | 0.958 | .. | .. |
| Age |  |  |  |  |  |  |
| <25 | 117 (43.8) | 150 (56.2) | 2.41 (1.73, 3.38) | <0.001 | 1.35 (0.89, 2.06) | 0.160 |
| 25–44 | 164 (31.5) | 357 (68.5) | 1.42 (1.06, 1.91) | 0.020 | 1.35 (1.00, 1.82) | 0.054 |
| 45–64 | 249 (30.6) | 566 (69.4) | 1.36 (1.03, 1.80) | 0.028 | 1.34 (1.01, 1.77) | 0.041 |
| ≥65 | 94 (24.4) | 291 (75.6) | 1.00 |  | 1.00 |  |
| Occupation |  |  |  |  |  |  |
| Farmer | 475 (28.4) | 1199 (71.6) | 0.56 (0.42, 0.76) | <0.001 | 0.60 (0.44, 0.82) | 0.001 |
| Students | 68 (57.6) | 50 (42.4) | 1.93 (1.22, 3.07) | 0.005 | 2.11 (1.20, 3.71) | 0.010 |
| Others | 81 (41.3) | 115 (58.7) | 1.00 |  | 1.00 |  |
| History of tuberculosis |  |  |  |  |  |  |
| New | 582 (31.6) | 1257 (68.4) | 1.00 |  | .. |  |
| Retreated | 42 (28.2) | 107 (71.8) | 0.85 (0.59, 1.23) | 0.382 | .. | .. |
| Diagnostic delay |  |  |  |  |  |  |
| <2 weeks | 157 (29.6) | 374 (70.4) | 1.00 |  | .. |  |
| 2–4 weeks | 124 (31.9) | 265 (68.1) | 1.11 (0.84, 1.48) | 0.452 | .. | .. |
| 4–8 weeks | 188 (32.5) | 391 (67.5) | 1.15 (0.89, 1.48) | 0.297 | .. | .. |
| ≥8 weeks | 155 (31.7) | 334 (68.3) | 1.11 (0.85, 1.44) | 0.461 | .. | .. |
| Chest cavitation |  |  |  |  |  |  |
| No | 385 (29.7) | 911 (70.3) | 1.00 |  | 1.00 |  |
| Yes | 239 (34.5) | 453 (65.5) | 1.25 (1.03, 1.52) | 0.027 | 1.22 (1.00, 1.50) | 0.054 |
| Sputum smear status |  |  |  |  |  |  |
| Negative | 258 (29.4) | 620 (70.6) | 1.00 |  | 1.00 |  |
| Positive | 366 (33.0) | 744 (67.0) | 1.18 (0.98, 1.43) | 0.087 | 1.19 (0.98, 1.46) | 0.085 |
| Drug-resistance profile |  |  |  |  |  |  |
| Pan-susceptible | 547 (31.9) | 1169 (68.1) | 1.00 |  | 1.00 |  |
| Other DR | 43 (25.3) | 127 (74.7) | 0.72 (0.50, 1.04) | 0.079 | 0.68 (0.47, 0.98) | 0.040 |
| MDR | 34 (33.3) | 68 (66.7) | 1.07 (0.70, 1.63) | 0.759 | 1.00 (0.65, 1.54) | 1.000 |
| Beijing strain |  |  |  |  |  |  |
| No | 158 (25.6) | 458 (74.4) | 1.00 |  | 1.00 |  |
| Yes | 466 (34.0) | 906 (66.0) | 1.49 (1.21, 1.84) | <0.001 | 1.46 (1.17, 1.82) | 0.001 |

Data are n (%) unless otherwise specified. OR=odds ratio. aOR=adjusted odds ratio. DR=drug resistance. MDR=multidrug resistance.

## Discussion

To our knowledge, this was the longest longitudinal population-based genomic epidemiological study of TB transmission in rural China. The overall cumulative clustering rate during the study period was 31·4%. Among 196 genomic-clustered patients with confirmed epidemiological links, 32·7% of transmission occurred between family contacts and 67·3% between social contacts. Of all MDR-TB cases diagnosed during the study, 81.4% were likely from transmission of MDR strains. The average time interval between the diagnosis of clustered student contacts was much shorter than for non-student contacts.

Close contacts of TB patients are at a high-risk for TB transmission. A recent systematic review suggested that the pooled prevalence of TB in close contacts was 3·6% (95% CI: 3·3-4·0), and that contact investigation could increase TB case notification and thereby decrease the incidence of TB in the population.^18^ In developed countries, index cases have been found to have 6-12 close contacts^19-22^ who could potentially constitute 10-20%^23,24^ of the total TB burden. Accordingly, the WHO strongly recommends systematic screening for TB disease among close contacts.^5,25^ In China, however, the importance of screening close contacts has been underappreciated and its role in case finding has been limited. In China, only 2-3 close contacts per index case were identified and most of these were family members, and it was therefore estimated that close contacts would contribute only about 1·0-3·3% of the total TB patients.^26,27^ A recent community-based study of active case-finding that included 320,000 people in 10 provinces in China during 2013-2015 concluded that screening of close contacts contributed less then 2% of TB patients.^7^

Our study found that 41·8% of clustered patients were close contacts of other patients and contributed 9·9% of the total TB patients, suggesting a high risk of TB transmission among close contacts in rural China. However, it was only through in-depth investigation of these genomic-clustered patients that we could identify the epidemiological links that are usually overlooked in routine contact investigation. In contrast to our findings, the routine TB contact investigations that were conducted in the study sites during the duration of our study registered only 2·4 close contacts per index case who contributed just 1·8% of the total patients. Without the WGS data that identified clustered patients and directed our investigations, nearly 80% of the transmission events between close contacts would have been missed. Most secondary cases were diagnosed between 1-3 years after the index case, but the long study duration of 12 years, using WGS to analyze all TB isolates in the study areas, allowed us to identify clustered isolates that would be missed in studies with a shorter duration.

Screening of close contacts in China has been poorly implemented for a number of reasons. The huge number of TB patients, limited resources for TB prevention and control and the stigma of tuberculosis all contribute to a failure to identify and screen many close contacts. When contact screening is poorly implemented, the contribution of close contacts to the overall TB burden is underestimated and therefore less attention is paid to the screening, creating a vicious cycle. WHO guidelines defined close contacts as persons sharing an enclosed space with the index case during the three months prior to commencement of the current treatment episode.^25^ While the identification of family contacts is generally straightforward, the definition and identification of non-family contacts is difficult. In China, non-family contacts refer mainly to classmates and colleagues.^27^ However, in this study we found that many patients with confirmed epidemiologic links were contacts either within the community or contacts who shared the aggregated settings outside the community. Therefore, in addition to family contacts, we classified others as social contacts in the hope of understanding where contacts occur and highlighting their relevance for TB transmission in rural China.

We found that 67·3% of the transmission occurred among social contacts in rural China, which was significantly higher than has been found in Vietnam, Zambia and South Africa (15-50%).^28,29^ This suggests that active case-finding in rural China should go beyond family contacts to include social contacts. In this study we proposed three categories of social contacts: relatives outside of the immediate family; people who know each other and live in the same community or village; and people frequenting aggregated settings outside the community. Largely due to differences in climate and lifestyle, the importance of the different categories of contacts varied between the two study sites. The villages in Wuchang are very small, consisting of perhaps only a dozen families and the houses are generally adjacent to each other. Wuchang is located in the northeast of China and has an average annual temperature of only 4°C. Consequently, villagers spend more than half of the year indoors and frequently interact to socialize and play cards with other villagers in the village shops. It is therefore not surprising that 51·2% of epidemiological links were attributable to transmission between social contacts within these communities. Wusheng, in contrast, is located in the southwest of China with an average annual temperature of 18°C. and villagers live in homes that are relatively scattered in the region. We found that transmission between villagers in this setting was less common than transmission at aggregated settings such as schools and teahouses, where individuals from several different villages congregate. Therefore, strategies for screening and social contact tracing must be tailored to the conditions and association habits of the local populations.

The purpose of active case-finding is to diagnose patients early and reduce transmission. Importantly, we found that the time interval between TB diagnosis of student patients and contacts was 3-4 times shorter than for family or community contacts (Figure 4B). Students are a special group in China and receive special attention from the government and society. Once a student patient is diagnosed with TB, their classmates and even schoolmates will be screened. If a similar enhanced screening strategy could be extended to other social contacts, patients might be diagnosed earlier in the course of their disease, thereby reducing rural TB transmission.

This study have several limitations. The clustering rate we calculated is likely an underestimation of the extent of local TB transmission. We previously found that the current passive case-finding strategy in Wusheng identified only about 30% of the incident TB patients^30^, but we were unable to even estimate the number of TB patients in the sampling region who were undiagnosed or diagnosed elsewhere. Some of the non-clustered patients could have been clustered with locally diagnosed TB patients without cultured isolates or patients diagnosed outside the study period or the geographic regions sampled. In addition, some clustered patients died or were lost to follow-up before the epidemiological investigations could be completed.

In conclusion, this long-term genomic-epidemiological study helped to delineate the patterns of TB transmission in rural China. Transmission appears to occur principally among close contacts, but contact investigation should be extended to include the social interactions that are common in the targeted population. Further improvement and implementation of screening of close contacts are critical to halt transmission and improve TB control in rural areas.

## Data Availability

All data produced in the present study are available upon reasonable request to the authors.

## Contributors

ML, QJ, CY, LX, FL, CC and QG conceived, designed and managed the study. SZ (Sheng Zhong), XS, YP, SZ (Shu Zhang), MG, and YQ contributed the demographic, clinical and microbiological data. ML, MG, YP, SZ (Sheng Zhong), XS, and PM contributed to the epidemiological investigations. ML, QM, QJ, and PM did the data analyses. ML, QJ, CY, HT, and QG drafted and revised the manuscript. All coauthors reviewed and approved the final manuscript before submission.

## Acknowledgments

This study was funded by the National Science and Technology Major Project of China (grants 2017ZX10201302-006). It was also funded by Natural Science Foundation of China (81661128043) and Science and Technology Major Project of Shanghai. We thank the tuberculosis public health teams in the Wusheng County Center for Disease Control and Prevention (Wusheng CDC) and the Wuchang City Center for Tuberculosis Control and Prevention (Wuchang CTC).

## Data sharing

De-identified participant data from the study will be made available with publication to medical researchers on a not for profit basis by email request to the corresponding author for the purposes of propensity matching or meta-analysis. Detailed supplementary materials will be made available by email request to the corresponding author.

---

**Table S1.**
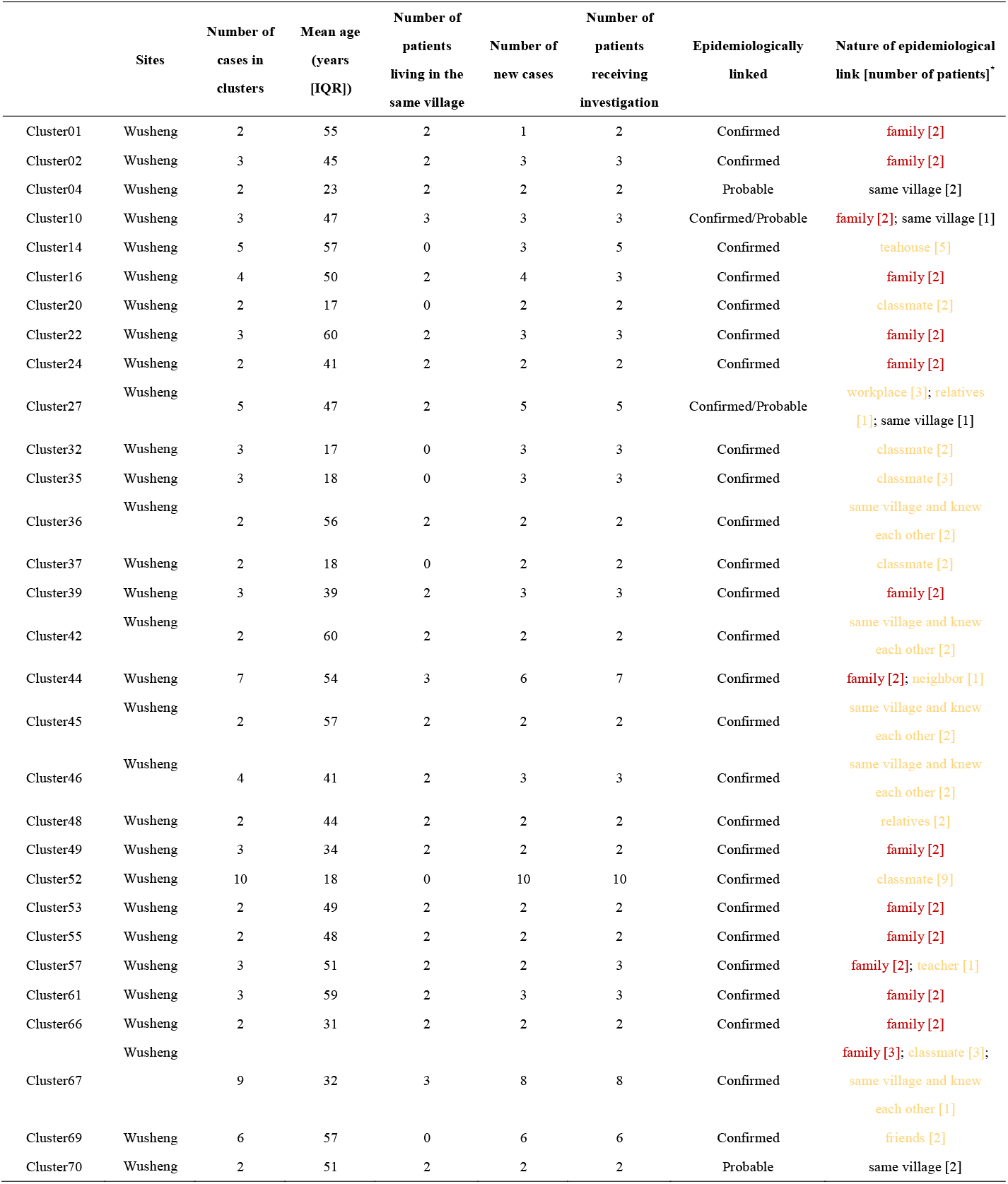
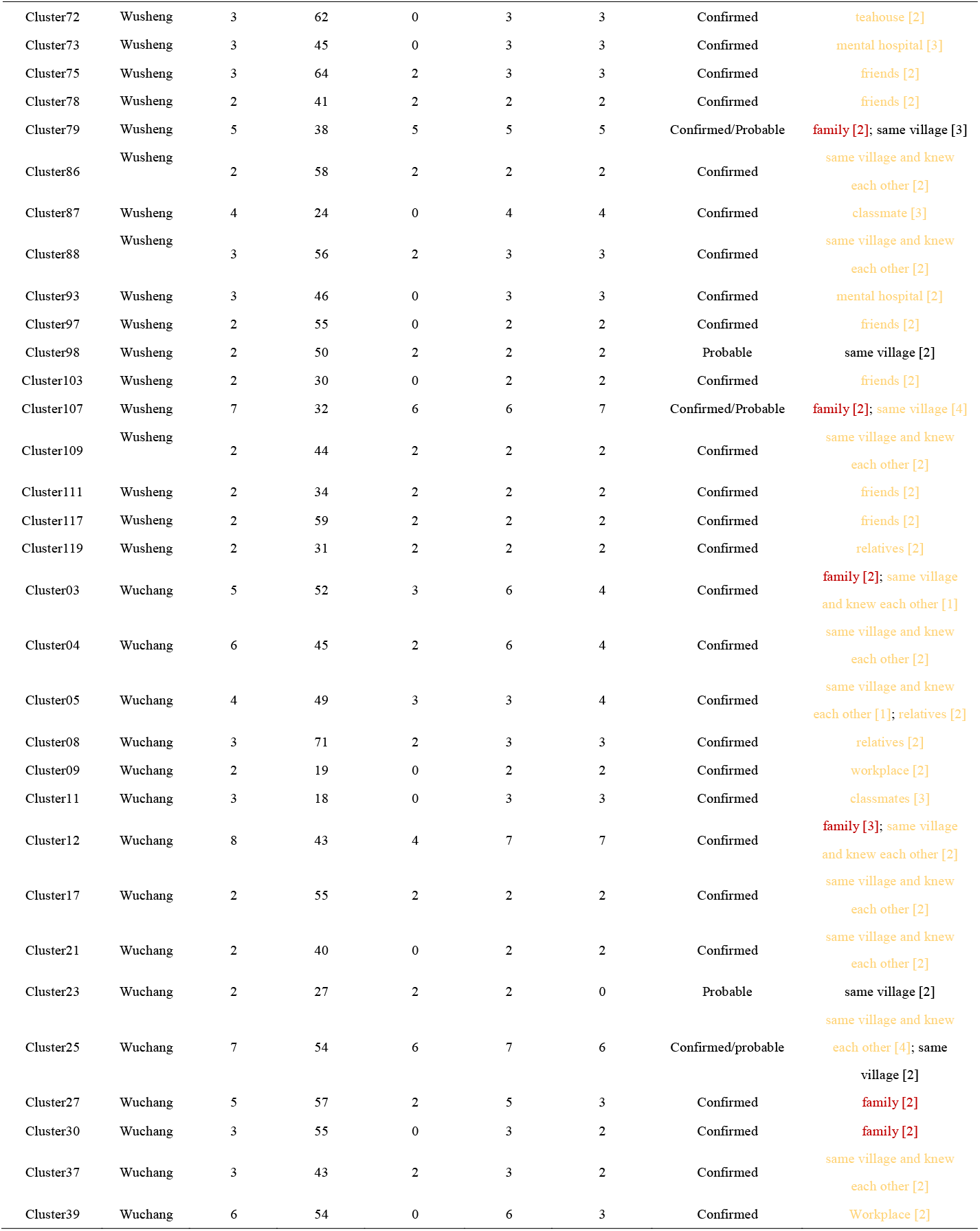
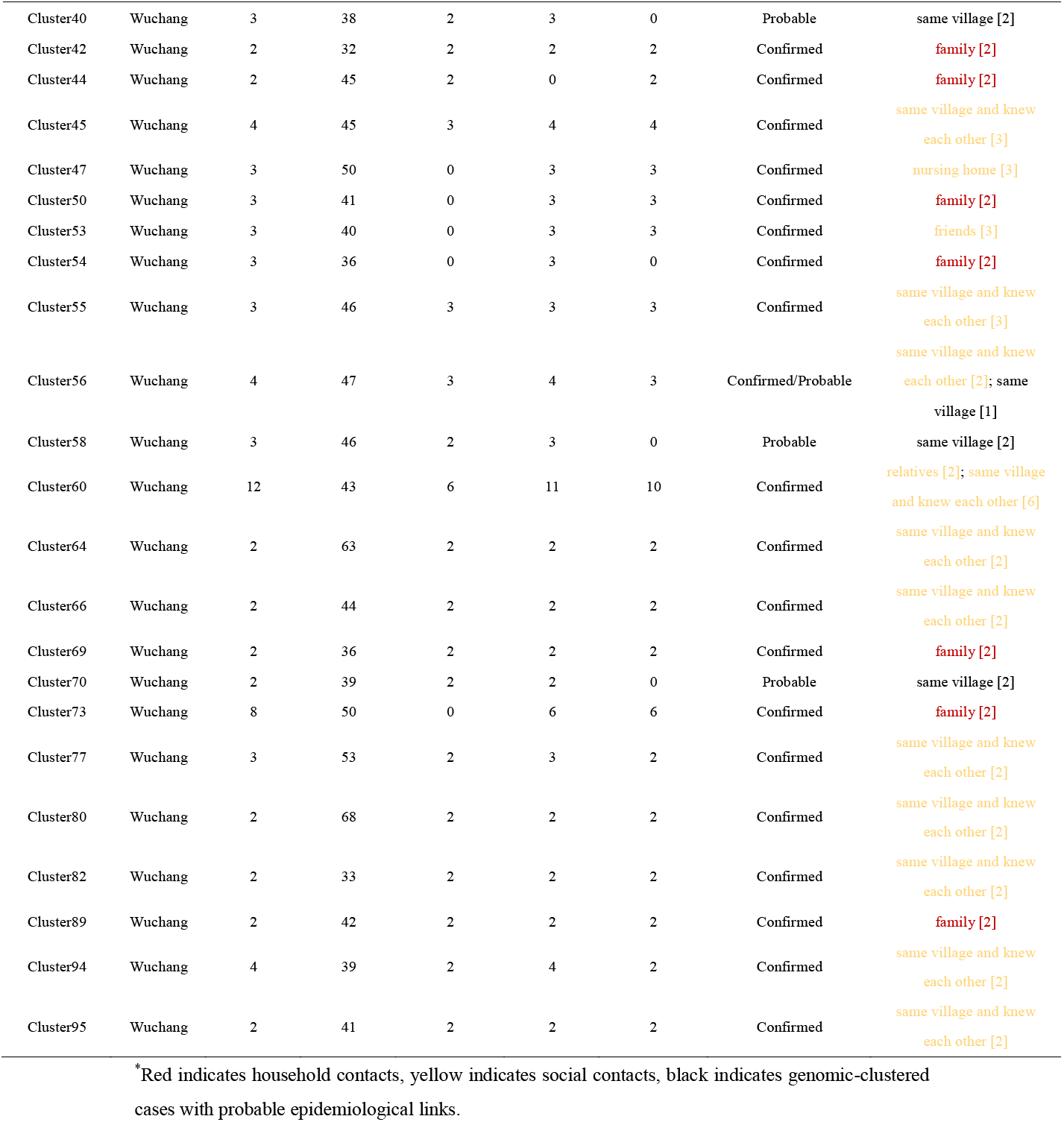
Characteristics of genomic clusters with confirmed or probable epidemiological links based on whole-genome sequencing analysis in Wusheng and Wuchang.

## FIGURE LEGENDS

**Figure S1.**
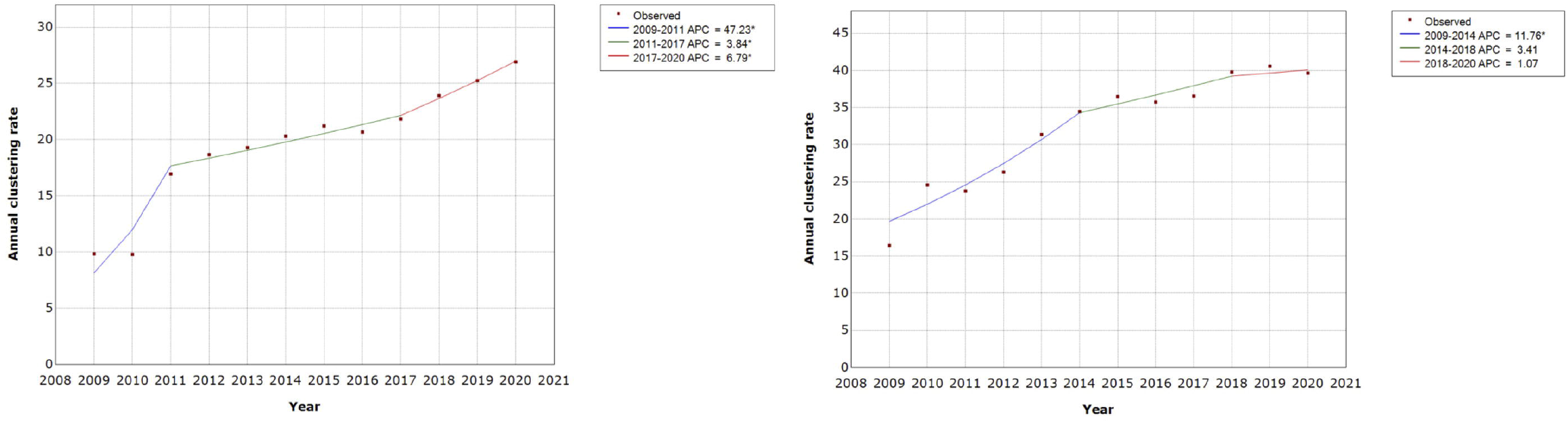
Changes in temporal trends in cumulative clustering rate in Wusheng (left) and Wuchang (right), 2009-2020. ^*^APC is significantly different from zero at the level of α=0.05.

**Figure S2.**
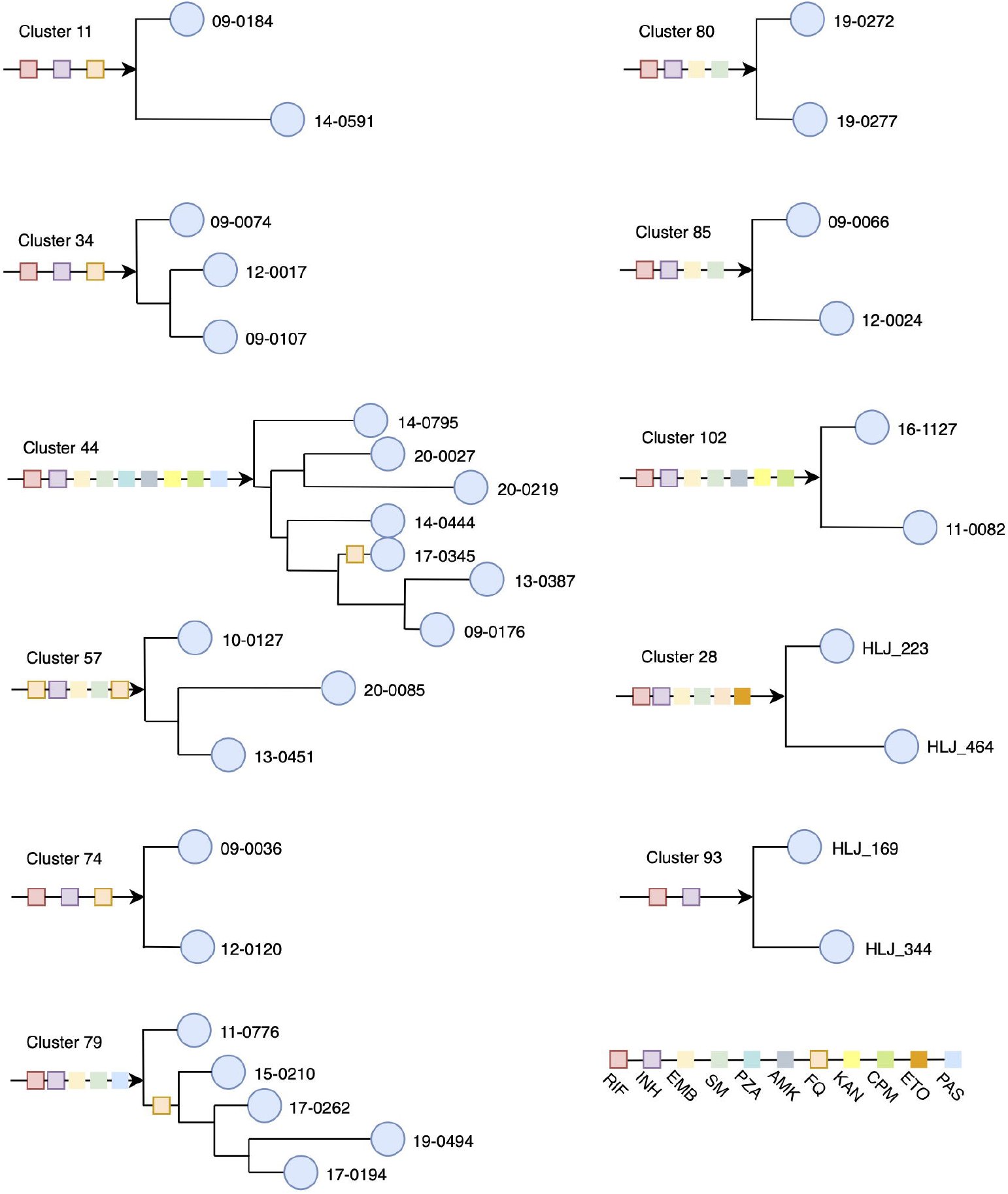
Transmission networks of multidrug-resistant tuberculosis based on genomic phylogeny and drug-resistance mutations. For each network, the arrow indicates the root and circles represent *Mycobacterium tuberculosis* isolates. M tuberculosis isolates are separated by lines with length representing genetic distance. Different colors of rectangle indicate resistance to different drugs. Two new MDR-TB patients were clustered with non-MDR-TB patients and were not included in the analysis.

